# *SELP* Asp603Asn and severe thrombosis in COVID-19 males: implication for anti P-selectin monoclonal antibodies treatment

**DOI:** 10.1101/2021.05.25.21257803

**Authors:** Chiara Fallerini, Sergio Daga, Elisa Benetti, Nicola Picchiotti, Kristina Zguro, Francesca Catapano, Virginia Baroni, Simone Lanini, Alessandro Bucalossi, Giuseppe Marotta, Margherita Baldassarri, Francesca Fava, Giada Beligni, Laura Di Sarno, Diana Alaverdian, Maria Palmieri, Susanna Croci, Andrea M. Isidori, Simone Furini, Elisa Frullanti, GEN-COVID Multicenter Study, Alessandra Renieri, Francesca Mari

## Abstract

Thromboembolism is a frequent cause of severity and mortality in COVID-19. However, the etiology of this phenomenon is not well understood. A cohort of 1,186 subjects, from the GEN-COVID consortium, infected by SARS-CoV-2 with different severity were stratified by sex and adjusted by age. Then, common coding variants from whole exome sequencing were mined by LASSO logistic regression. The homozygosity of the cell adhesion molecule P-selectin gene (*SELP)* rs6127 (c.1807G>A; p.Asp603Asn) which increases platelet activation is found to be associated with severity in the male subcohort of 513 subjects (Odds Ratio= 2.27, 95% Confidence Interval 1.54-3.36). As the *SELP* gene is downregulated by testosterone, the odd ratio is increased in males older than 50 (OR 2.42, 95% CI 1.53-3.82). Asn/Asn homozygotes have increased D-dimers values especially when associated with poly Q≥23 in the androgen receptor (*AR)* gene (OR 3.26, 95% CI 1.41-7.52). These results provide a rationale for the repurposing of antibodies against P-selectin as adjuvant therapy in rs6127 male homozygotes especially if older than 50 or with impaired *AR* gene.

**Key points:**

○ The functional polymorphism rs6127 (p.Asp603Asn) in the testosterone-regulated *SELP* gene associates with COVID-19 severity and thrombosis.
○ Conditions with decreased testosterone (old males), or decreased testosterone efficacy (*AR* gene polyQ ≥ 23) strengthen the association.

## INTRODUCTION

Since its onset in early 2020, the global pandemic of coronavirus disease 2019 (COVID-19), caused by SARS-CoV-2, has posed a multitude of challenges and triggered intensive research on the disease and the virus. However, a lot of questions still remain unanswered, especially those regarding its heterogeneity of clinical manifestations and the appropriate therapeutic strategies.

It is now widely recognized that COVID-19 is a systemic disease^1^ characterized by dysregulation of the immune system and presence of a hypercoagulable state^2^. COVID-19 severe patients show coagulation abnormalities with increased incidence of arterial thrombosis and mortality has been shown to correlate with elevated levels of both interleukin-6 (IL-6) and D-dimer^2,3^. These values have often been reported accompanied by a relatively modest decrease in platelet count^4^, and prolongation of the prothrombin time^3^.

Genetic bases of this prothrombotic susceptibility remain until now elusive, despite the fact that it is evident that phenotypic variability associated with the viral infection is obviously also due to host genetic factors. Some rare variants of genes involved in adaptive immunity have been identified by us and others in Mendelian forms of COVID-19^5-7^. Among common genetic factors, the protective role of the zero blood group has been linked to destabilization of the von Willebrand factor and protection from thrombosis^8^. We have shown that longer polyQ repeats (≥ 23) in the androgen receptor gene (*AR*) predisposes to severe COVID-19 outcome due to reduced anti-inflammatory and anti-thrombotic effect of testosterone^9^.

The testosterone inhibited P-selectin (*SELP*) gene encodes a cell adhesion molecule that is stored in the alpha-granules of platelets and in Weibel-Palade bodies of endothelial cells. It mediates the interaction of activated platelets on endothelium with leukocytes playing a key role in the thrombotic process^10,11^. A significantly increased P-selectin concentration in plasma samples of severe COVID-19 patients compared to healthy controls, together with other prothrombotic biomarkers has been recently demonstrated^12,13^.

Among *SELP* variants, the Asp603Asn functional polymorphism (rs6127; c.1807G>A) - previously reported as Asp562Asn or Asp541Asn - has been associated with risk of thrombotic disease, tissue damage in diabetes, abortion, as well as myocardial infarction^14-18^. The polymorphism is located within the consensus repeat domain of the SELP protein^19^, and it has been shown to affect the binding of P-selectin to its ligand (PSGL-1) on leukocytes, resulting in a protein that is more efficient at recruiting leukocytes to the endothelium^16^.

Here, we applied Ordered Logistic Regression (OLR) analysis on clinical data and LASSO logistic regression on WES data within our prospectively recruited GEN-COVID cohort and identified SELP as a key player for severity and thromboembolism in severe COVID-19.

## MATERIAL AND METHODS

A cohort of 1,186 SARS-CoV-2-infected subjects was recruited during the first pandemic wave within the Italian GEN-COVID Study (https://sites.google.com/dbm.unisi.it/gen-covid)^1^. ORL model was applied to the clinical WHO gradings, stratified by sex and adjusted by age, as described elsewhere^20^. The male subset consists of 513 COVID-19 male patients: 236 severe COVID-19 patients (cases) and 277 SARS-CoV-2 PCR-positive oligo-asymptomatic subjects (controls). WES with at least 97% coverage at 20x was performed using the NovaSeq6000 System (Illumina, San Diego, CA, USA) as previously described^1^. WES data were represented in a binary mode on a gene-by-gene basis^20^. LASSO logistic regression model was applied to a synthetic boolean representation under recessive model of genetic combinations ^20^. Association was confirmed by Chi Square Test. Longitudinal laboratory values were represented by linear graphs. Differences were assessed by Mann–Whitney test and displayed by box plots. Statistical procedures were carried out using R packages.

## RESULTS AND DISCUSSION

Within the Italian GEN-COVID cohort, we tested different combination of coding polymorphisms of human genes at homozygous state and found that the Asp603Asn polymorphism of the *SELP* gene correlate with severity in the subcohort of males (**Figure 1, panel A**) ^1,20^. The genotypic frequencies of the polymorphism in cases and controls were confirmed to be in Hardy-Weinberg equilibrium; the minor allele frequency in our cohort was similar to that reported in the European (non-Finnish) population in the gnomAD database (16.4% versus 15.9%) (https://gnomad.broadinstitute.org/).

**Figure 1.**
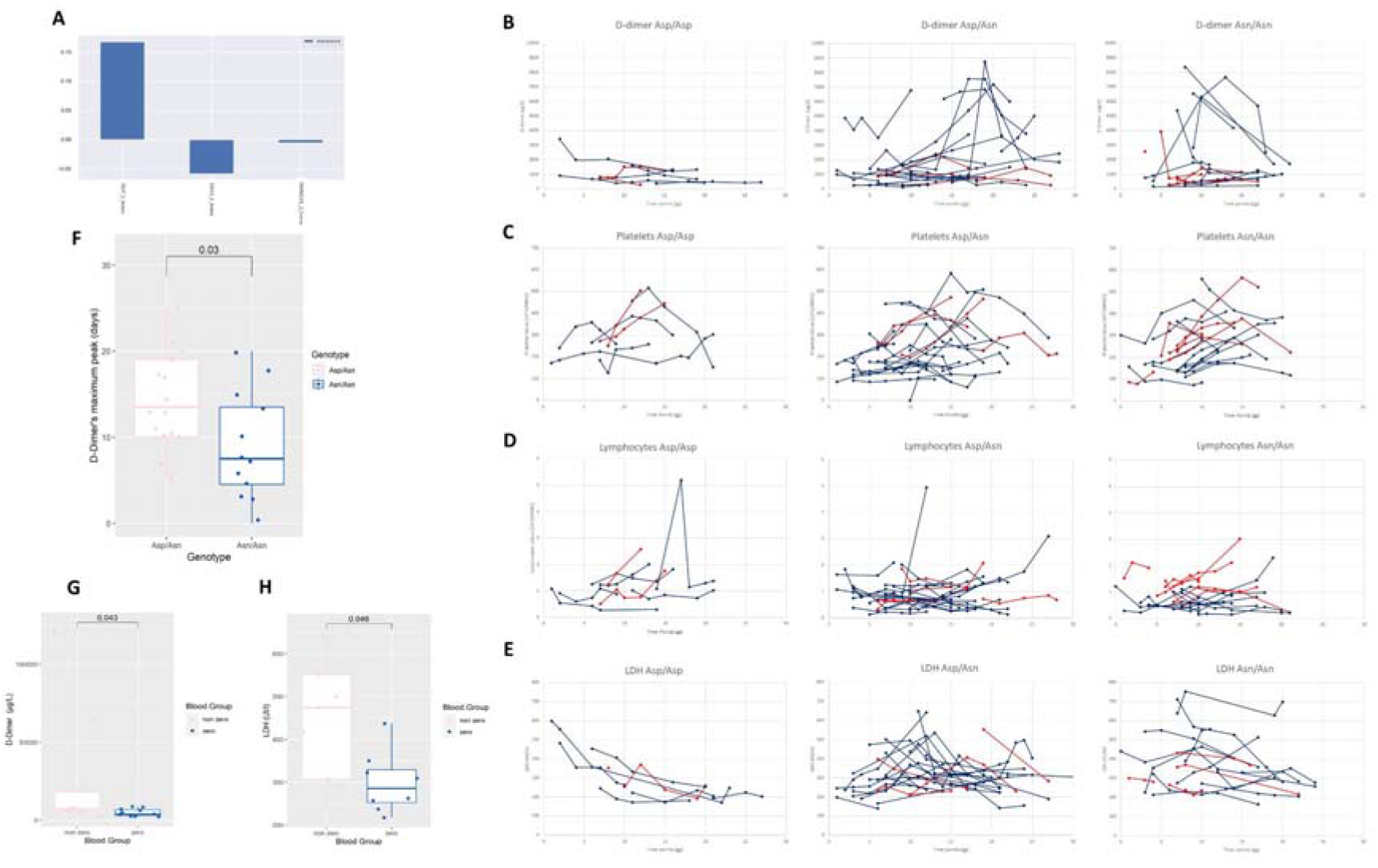
Homozygous genotype Asn/Asn at the polymorphic locus Asp603Asn (rs6127) is related to severity and to D-Dimer pick. **Panel A**. Selection of *SELP* gene as relevant for severity. LASSO logistic regression on boolean representation of homozygous common bi-allelic polymorphism of autosomal genes in males is presented (see paper Picchiotti N. et al. 2021 for complete representations)^21^. The LASSO logistic regression model provides an embedded feature selection method within the binary classification tasks (cases vs controls). The upward histogram means positive weights, i.e the specific variant at the specific locus (feature) contribute to severity of COVID-19. *SELP*_1_homo = homozygous genotype Asn/Asn at the polymorphic locus Asp603Asn (rs6127). The downward histograms means negative weights, contributing to mildness of COVID-19. *COG3*_1_homo = homozygous genotype Ser/Ser at the polymorphic locus Leu825Ser (rs3014902). *COG3* gene encodes for a vesicle docking protein involved in viral trafficking. *TMEM221*_2_homo = homozygous genotype Ala/Ala at the polymorphic locus Thr66Ala (rs4808641). *TMEM221* gene encodes for a transmembrane protein. **Panel B-E**. Longitudinal laboratory data related to thrombosis and severity. Linear graphs of 4 laboratory values: D-dimer μg/L (**Panel B**), platelets 10^3/mmc (**Panel C**), lymphocytes 10^3/mmc (**Panel D**), LDH UI/L (**Panel E**). As expected the Asn/Asn homozygous genotype was over-represented (36.53%). Values are reported on the Y-axis. In each graph the time point “0” (X-axis) represents the day of onset of COVID-19 symptoms. Each line represents each severe hospitalized patient (see methods). Each point represents the different time point (day) in which the different values have been measured. Patients aged ≥55 years are indicated in blue while patients aged <55 years in red. From left to right patients having Asp/Asp homozygous; Asp/Asn heterozygous; and Asn/Asn homozygous genotype. Older patients only (blue) and Asp/Asn-Asn/Asn genotype only show the D-dimer pick. Accordingly, older patients of these 2 genotypes have more platelet consumption and higher LDH values. **Panel F**. The D-dimer pick is earlier in the Asn/Asn (median = 7.5 days) than the Asp/Asn genotype (p= 3×10^−2^ by Mann Whitney test). Box plot of patients with D-dimer values above 2000 µg/l were represented. Only Asp/Asn (light blue) and Asn/Asn (pink) genotypes are represented because patients with Asp/Asp genotype do not have the pick and do not show value above 2.000. **Panel G-H**. The non-zero group associates with higher D-dimer (**Panel G**) and LDH values (**Panel H**). Severe hospitalized patients with zero blood group =light blue; non-zero blood group =pink in box plots.

We reasoned that the hyper-inflammatory and hyper-thrombotic state, due to viral injury of the vascular endothelium, leads to the release of P-selectin by activated platelets, driving thrombosis and vascular inflammation especially in those individuals with enhanced P-selectin activities due a double copy of Asparagine in position 603 without any other additional coding polymorphisms^16^. These results are in line with the demonstration that SARS-CoV-2 induces thrombosis by binding to ACE2 on platelets and subsequent integrin αIIbβ3 activation and P-selectin expression^21^, and that P-selectin soluble isoform is increased in thrombosis^10,11^ and severe COVID-19^12,13^.

The association between Asp603Asn polymorphism (rs6127) in homozygous state, in absence of other coding polymorphisms, and severity is quite strong with an OR 2.27 in males (95% CI, 1.54 to 3.36; p-value 2.8×10^−5^, **Table 1a**). We reasoned that, since *SELP* transcription is inhibited by androgens^22^, the strength of the association should increase with age. Indeed, in males aged ≥ 50 years we found an increased OR of 2.42 (95% CI, 1.53 to 3.82; p-value 1.19×10^−4^, **Table 1b**). While the significance is, instead, lost in young males (<50 years), (data not shown p= 1.1×10^−1^).

**Table 1.**
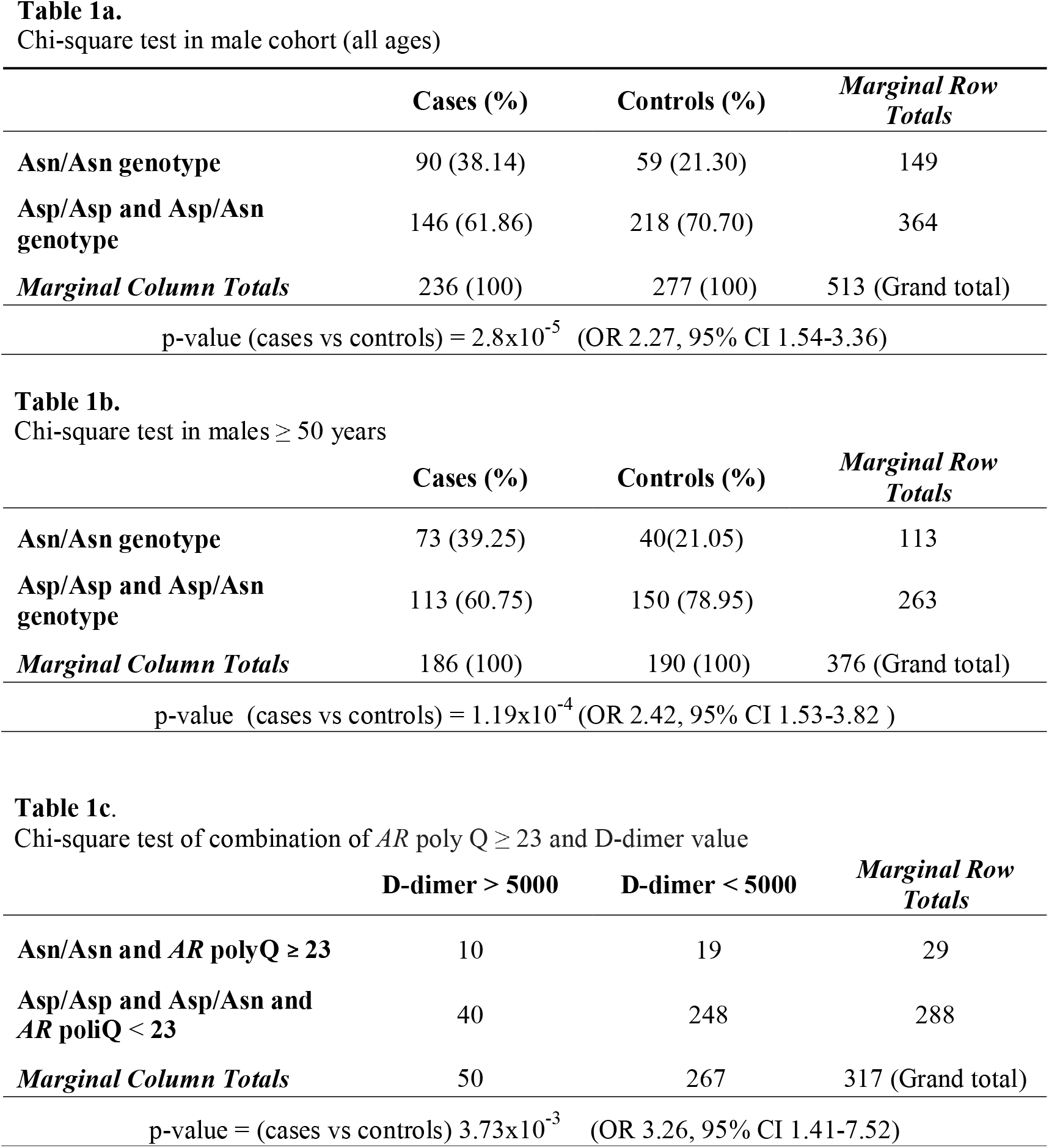
Chi-square test in male cohort calculated for all ages **(Table 1a**); for age ≥ 50 years (**Table 1b**); and combination of *AR* poly-Q ≥ 23 and D-dimer value (**Table 1c**).

In a subset of 52 severely affected hospitalised males, 4 main laboratory parameters indicating thrombosis (D-dimer, platelets and LDH) and severity (lymphocytes) were longitudinally followed (**Figure 1, Panel B-E**). We observed that the maximum pick, over 10 times of the normal value, was exclusive of Asp/Asn and Asn/Asn genotypes and older patients (**Figure 1, Panel B-E)**. The pick timing was earlier in Asn/Asn (median 7.5 days from infection) than Asp/Asn (median 13.5 days from infection), (p-value = 3×10^−2^, **Figure 1, panel F**). As the von Willebrand factor (vWF) is a downstream effector for clotting, the non-zero blood group, associating with more stable vWF, also correlate with higher D-dimer and LDH values (**Figure 1, panel G-H)**, in agreement with previous reports^8^.

Given the higher association of the *SELP* polymorphism in older males we reasoned that the *AR* poly-Q status would impact on the *SELP* genotype^9^: the combination of poly-Q polymorphism ≥ 23 with homozygous *SELP* polymorphism versus D-dimer value reached an OR of 3.26 (95% CI 1.41-7.52; p-value 3.73×10^−3^), (**Table 1c**). This result indicates that the two polymorphisms enhance each other, and that the *SELP* Asp603Asn and the *AR* poly-Q are two factors belonging to the same puzzle contributing to thrombosis in COVID-19 in males.

Anti-*P-Selectin* monoclonal antibodies have been developed for human use: the phase 3 Inclacumab and the FDA and EMA approved Crizanlizumab. The latter is indicated to prevent vaso-occlusive crises in patients with sickle cell disease and to decrease inflammation, blocking leucocyte and platelet adherence to the vessel wall^23-25^. A general clinical trial to test the efficacy and safety of Crizanlizumab in patients hospitalized with COVID-19 is ongoing (https://clinicaltrials.gov/ct2/show/study/NCT04435184), in not selected patients. Clinical trials in COVID-19 hospitalised males with *SELP* rs6127 c.1807G>A p.Asp603Asn should be encouraged.

In conclusion, we identified *SELP* rs6127 polymorphism as the elusive genetic factor predisposing COVID-19 patients to thromboembolism leading to life-threatening disease. We showed that predisposition increases if the protective effect of testosterone is lost either by age or because of additional genetic factors such as poly Q≥23 in the *AR* gene. This knowledge provides a rationale for repurposing anti P-selectin monoclonal antibodies as personalized adjuvant therapy in men affected by COVID-19.

## Data Availability

The data and samples referenced here are housed in the GEN-COVID Patient Registry and the GEN-COVID Biobank and are available for consultation. You may contact the corresponding author, Prof. Alessandra Renieri.

## ACKNOWLEDGMENT

This study is part of the GEN-COVID Multicenter Study, https://sites.google.com/dbm.unisi.it/gen-covid, the Italian multicenter study aimed at identifying the COVID-19 host genetic bases. Specimens were provided by the COVID-19 Biobank of Siena, which is part of the Genetic Biobank of Siena, member of BBMRI-IT, of Telethon Network of Genetic Biobanks (project no. GTB18001), of EuroBioBank, and of RDConnect. We thank the CINECA consortium for providing computational resources and the Network for Italian Genomes (NIG) http://www.nig.cineca.it for its support. We thank private donors for the support provided to A.R. (Department of Medical Biotechnologies, University of Siena) for the COVID-19 host genetics research project (D.L n.18 of March 17, 2020). We also thank the COVID-19 Host Genetics Initiative (https://www.covid19hg.org/), MIUR project “Dipartimenti di Eccellenza 2018-2020” to the Department of Medical Biotechnologies University of Siena, Italy and “Bando Ricerca COVID-19 Toscana’’ project to Azienda Ospedaliero Universitaria Senese. We also thank Intesa SanPaolo for the 2020 charity fund dedicated to the project “N. B/2020/0119 Identificazione delle basi genetiche determinanti la variabilità clinica della risposta a COVID-19 nella popolazione italiana’’; the Italian Ministry of University and Research for funding within the “Bando FISR 2020” in COVID-19 and and the Istituto Buddista Italiano Soka Gakkai for funding the project “PAT-COVID: Host genetics and pathogenetic mechanisms of COVID-19” (ID n. 2020-2016_RIC_3).

## AUTHORSHIP

### Contribution

AR, FM and EF designed the study; CF, SD, EB, NP, KZ, FC, VB, GB, LDS, DA, SL, SC, MP, AB, GM, AMI, EF, SF analysed the data; EB, KZ, NP, SF performed statistical analysis; MB, FF and GEN-COVID Multicenter Study provided clinical data; AR and FM supervised the study and all authors edited the manuscript and approved its final version.

## ETHICS APPROVAL

The study (GEN-COVID) was consistent with Institutional guidelines and approved by the University Hospital (Azienda Ospedaliero-Universitaria Senese) Ethical Review Board, Siena, Italy (Prot n. 16929, dated March 16, 2020).

## GEN-COVID Multicenter Study (https://sites.google.com/dbm.unisi.it/gen-covid)

Floriana Valentino^1,2^, Gabriella Doddato^1,2^, Annarita Giliberti^1,2^, Rossella Tita^7^, Sara Amitrano^7^, Mirella Bruttini^1,2,7^, Ilaria Meloni^1,2^, Anna Maria Pinto^7^, Maria Antonietta Mencarelli^7^, Caterina Lo Rizzo^7^, Francesca Montagnani^2,8^, Miriam Lucia Carriero^1,2^, Massimiliano Fabbiani^8^, Ilaria Rancan^8^, Barbara Rossetti^8^, Mario Tumbarello^2,8^, Elena Bargagli^9^, Laura Bergantini^9^, Miriana D’Alessandro^9^, Paolo Cameli^9^, David Bennett^9^, Federico Anedda^10^, Simona Marcantonio^10^, Sabino Scolletta^10^, Federico Franchi^10^, Maria Antonietta Mazzei^11^, Susanna Guerrini^11^, Edoardo Conticini^12^, Luca Cantarini^12^, Bruno Frediani^12^, Danilo Tacconi^13^, Chiara Spertilli Raffaelli^13^, Marco Feri^14^, Alice Donati^14^, Raffaele Scala^15^, Luca Guidelli^15^, Genni Spargi^16^, Marta Corridi^16^, Cesira Nencioni^17^, Leonardo Croci^17^, Gian Piero Caldarelli^18^, Maurizio Spagnesi^19^, Paolo Piacentini^19^, Maria Bandini^19^, Elena Desanctis^19^, Silvia Cappelli^19^, Anna Canaccini^20^, Agnese Verzuri^20^ Valentina Anemoli^20^, Agostino Ognibene^21^, Alessandro Pancrazi^21^, Maria Lorubbio^21^, Massimo Vaghi^22^, Antonella D’Arminio Monforte^23^, Esther Merlini^23^, Federica Gaia Miraglia^23^, Mario U. Mondelli^24,25^, Raffaele Bruno^24,25^, Marco Vecchia^24,^, Stefania Mantovani^24^, Serena Ludovisi^24,25^, Massimo Girardis^26^, Sophie Venturelli^26^, Stefano Busani^26^, Andrea Cossarizza^27^, Andrea Antinori^28^ Alessandra Vergori^28^, Arianna Emiliozzi^28^, Stefano Rusconi^29,30^, Matteo Siano^30^, Arianna Gabrieli^30^, Agostino Riva^29,30^, Daniela Francisci^31,32^, Elisabetta Schiaroli^31^, Francesco Paciosi^31^, Andrea Tommasi^31^, Pier Giorgio Scotton^33^, Francesca Andretta^33^, Sandro Panese^34^, Renzo Scaggiante^35^, Francesca Gatti^35^, Saverio Giuseppe Parisi^36^, Francesco Castelli^37^, Eugenia Quiros-Roldan^37^, Melania degli Antoni^37^, Isabella Zanella^38^, Matteo Della Monica^39^, Carmelo Piscopo^39^, Mario Capasso^40,41,42^, Roberta Russo^40,42^, Immacolata Andolfo^40,41^, Achille Iolascon^40,41^, Giuseppe Fiorentino^43^, Massimo Carella^44^, Marco Castori^44^, Filippo Aucella^45^, Pamela Raggi^46^, Carmen Marciano^46^, Rita Perna^46^, Matteo Bassetti^47,48^, Antonio Di Biagio^48^ Maurizio Sanguinetti^49,50^, Luca Masucci^49,50^, Serafina Valente^51^, Oreste De Vivo^51^, Marco Mandalà^52^, Alessia Giorli^52^, Lorenzo Salerni^52^, Patrizia Zucchi^53^, Pierpaolo Parravicini^53^, Elisabetta Menatti^54^, Stefano Baratti^55^, Tullio Trotta^56^, Ferdinando Giannattasio^56^, Gabriella Coiro^56^, Fabio Lena^57^, Domenico A. Coviello^58^, Cristina Mussini^59^, Giancarlo Bosio^60^, Enrico Martinelli^60^, Sandro Mancarella^61^, Luisa Tavecchia^61^, Mary Ann Belli^61^, Lia Crotti^62,63,64,65,66^, Gianfranco Parati^62,63^, Marco Gori^4,67^, Chiara Gabbi^68^, Maurizio Sanarico^69^, Stefano Ceri^70^, Pietro Pinoli^70^, Francesco Raimondi^71^, Filippo Biscarini^72^, Alessandra Stella^72^, Marco Rizzi^73^, Franco Maggiolo^73^, Diego Ripamonti^73^, Claudia Suardi^74^, Tiziana Bachetti^75^, Maria Teresa La Rovere^76^, Simona Sarzi-Braga^77^, Maurizio Bussotti^78^ Simona Dei^79^, Katia Capitani^2,80^, Sabrina Ravaglia^81^, Rosangela Artuso^82^, Antonio Perrella^83^, Francesco Bianchi^2,83^, Davide Romani^19^, Paola Bergomi^84^, Emanuele Catena^84^, Riccardo Colombo^84^, Sauro Luchi^85^, Giovanna Morelli^85^, Paola Petrocelli^85^, Valentina Perticaroli^1,2,7^, Mirjam Lista^1,2^, Silvia Baroni^86^, Francesco Vladimiro Segala^87^, Alessandra Guarnaccia^49^, Giuseppe Merla^40,88^, Gabriella Maria Squeo^88^

8. Dept of Specialized and Internal Medicine, Tropical and Infectious Diseases Unit, Azienda Ospedaliera Universitaria Senese, Siena, Italy

9. Unit of Respiratory Diseases and Lung Transplantation, Department of Internal and Specialist Medicine, University of Siena

10 Dept of Emergency and Urgency, Medicine, Surgery and Neurosciences, Unit of Intensive Care Medicine, Siena University Hospital, Italy

11. Department of Medical, Surgical and Neurosciences and Radiological Sciences, Unit of Diagnostic Imaging, University of Siena

12 Rheumatology Unit, Department of Medicine, Surgery and Neurosciences, University of Siena, Policlinico Le Scotte, Italy

13. Department of Specialized and Internal Medicine, Infectious Diseases Unit, San Donato Hospital Arezzo, Italy

14. Dept of Emergency, Anesthesia Unit, San Donato Hospital, Arezzo, Italy

15. Department of Specialized and Internal Medicine, Pneumology Unit and UTIP, San Donato Hospital, Arezzo, Italy

16. Department of Emergency, Anesthesia Unit, Misericordia Hospital, Grosseto, Italy

17. Department of Specialized and Internal Medicine, Infectious Diseases Unit, Misericordia Hospital, Grosseto, Italy

18. Laboratory Medicine Department, Misericordia Hospital, Grosseto, Italy

19. Department of Preventive Medicine, Azienda USL Toscana Sud Est, Italy

20. Territorial Scientific Technician Department, Azienda USL Toscana Sud Est, Italy

21. Laboratory Medicine Department, San Donato Hospital, Arezzo, Italy

22. Chirurgia Vascolare, Ospedale Maggiore di Crema, Italy

23. Department of Health Sciences, Clinic of Infectious Diseases, ASST Santi Paolo e Carlo, University of Milan, Italy

24. Division of Infectious Diseases and Immunology, Fondazione IRCCS Policlinico San Matteo, Pavia, Italy

25. Department of Internal Medicine and Therapeutics, University of Pavia, Italy

26. Department of Anesthesia and Intensive Care, University of Modena and Reggio Emilia, Modena, Italy

27. Department of Medical and Surgical Sciences for Children and Adults, University of Modena and Reggio Emilia, Modena, Italy

28. HIV/AIDS Department, National Institute for Infectious Diseases, IRCCS, Lazzaro Spallanzani, Rome, Italy

29. III Infectious Diseases Unit, ASST-FBF-Sacco, Milan, Italy

30. Department of Biomedical and Clinical Sciences Luigi Sacco, University of Milan, Milan, Italy

31. Infectious Diseases Clinic, Department of Medicine, Azienda Ospedaliera di Perugia and University of Perugia, Santa Maria Hospital, Perugia, Italy

32. Infectious Diseases Clinic, “Santa Maria” Hospital, University of Perugia, Perugia, Italy

33. Department of Infectious Diseases, Treviso Hospital, Local Health Unit 2 Marca Trevigiana, Treviso, Italy

34. Clinical Infectious Diseases, Mestre Hospital, Venezia, Italy.

35. Infectious Diseases Clinic, ULSS1, Belluno, Italy

36. Department of Molecular Medicine, University of Padova, Italy

37. Department of Infectious and Tropical Diseases, University of Brescia and ASST Spedali Civili Hospital, Brescia, Italy

38. Department of Molecular and Translational Medicine, University of Brescia, Italy; Clinical Chemistry Laboratory, Cytogenetics and Molecular Genetics Section, Diagnostic Department, ASST Spedali Civili di Brescia, Italy

39. Medical Genetics and Laboratory of Medical Genetics Unit, A.O.R.N. “Antonio Cardarelli”, Naples, Italy

40. Department of Molecular Medicine and Medical Biotechnology, University of Naples Federico II, Naples, Italy

41. CEINGE Biotecnologie Avanzate, Naples, Italy

42. IRCCS SDN, Naples, Italy

43. Unit of Respiratory Physiopathology, AORN dei Colli, Monaldi Hospital, Naples, Italy

44. Division of Medical Genetics, Fondazione IRCCS Casa Sollievo della Sofferenza Hospital, San Giovanni Rotondo, Italy

45. Department of Medical Sciences, Fondazione IRCCS Casa Sollievo della Sofferenza Hospital, San Giovanni Rotondo, Italy

46. Clinical Trial Office, Fondazione IRCCS Casa Sollievo della Sofferenza Hospital, San Giovanni Rotondo, Italy

47. Department of Health Sciences, University of Genova, Genova, Italy

48. Infectious Diseases Clinic, Policlinico San Martino Hospital, IRCCS for Cancer Research Genova, Italy

49. Microbiology, Fondazione Policlinico Universitario Agostino Gemelli IRCCS, Catholic University of Medicine, Rome, Italy

50. Department of Laboratory Sciences and Infectious Diseases, Fondazione Policlinico Universitario A. Gemelli IRCCS, Rome, Italy

51. Department of Cardiovascular Diseases, University of Siena, Siena, Italy

52. Otolaryngology Unit, University of Siena, Italy

53. Department of Internal Medicine, ASST Valtellina e Alto Lario, Sondrio, Italy

54. Study Coordinator Oncologia Medica e Ufficio Flussi, Sondrio, Italy

55. Department of Infectious and Tropical Diseases, University of Padova, Padova, Italy

56. First Aid Department, Luigi Curto Hospital, Polla, Salerno, Italy

57. Local Health Unit-Pharmaceutical Department of Grosseto, Toscana Sud Est Local Health Unit, Grosseto, Italy

58. U.O.C. Laboratorio di Genetica Umana, IRCCS Istituto G. Gaslini, Genova, Italy.

59. Infectious Diseases Clinics, University of Modena and Reggio Emilia, Modena, Italy.

60. Department of Respiratory Diseases, Azienda Ospedaliera di Cremona, Cremona, Italy

61. U.O.C. Medicina, ASST Nord Milano, Ospedale Bassini, Cinisello Balsamo (MI), Italy

62. Istituto Auxologico Italiano, IRCCS, Department of Cardiovascular, Neural and Metabolic Sciences, San Luca Hospital, Milan, Italy.

63. Department of Medicine and Surgery, University of Milano-Bicocca, Milan, Italy

64. Istituto Auxologico Italiano, IRCCS, Center for Cardiac Arrhythmias of Genetic Origin, Milan, Italy.

65. Istituto Auxologico Italiano, IRCCS, Laboratory of Cardiovascular Genetics, Milan, Italy.

66. Member of the European Reference Network for Rare, Low Prevalence and Complex Diseases of the Heart-ERN GUARD-Heart

67. University Cote d’Azur, Inria, CNRS, I3S, Maasai

68. Independent Medical Scientist, Milan, Italy

69. Independent Data Scientist, Milan, Italy

70. Department of Electronics, Information and Bioengineering (DEIB), Politecnico di Milano, Milano, Italy

71. Scuola Normale Superiore, Pisa, Italy

72. CNR-Consiglio Nazionale delle Ricerche, Istituto di Biologia e Biotecnologia Agraria (IBBA), Milano, Italy.

73. Unit of Infectious Diseases, ASST Papa Giovanni XXIII Hospital, Bergamo, Italy

74. Fondazione per la ricerca Ospedale di Bergamo, Bergamo, Italy

75. Direzione Scientifica, Istituti Clinici Scientifici Maugeri IRCCS, Pavia, Italy.

76. Istituti Clinici Scientifici Maugeri IRCCS, Department of Cardiology, Institute of Montescano, Pavia, Italy.

77. Istituti Clinici Scientifici Maugeri, IRCCS, Department of Cardiac Rehabilitation, Institute of Tradate (VA), Italy.

78. Cardiac Rehabilitation Unit, Fondazione Salvatore Maugeri, IRCCS, Scientific Institute of Milan, Milan, Italy.

79. Health Management, Azienda USL Toscana Sudest, Tuscany, Italy.

80. Core Research Laboratory, ISPRO, Florence, Italy

81. IRCCS C. Mondino Foundation, Pavia, Italy

82. Medical Genetics Unit, Meyer Children’s University Hospital, Florence, Italy

83. Department of Medicine, Pneumology Unit, Misericordia Hospital, Grosseto, Italy.

84. Department of Anesthesia and Intensive Care Unit, ASST Fatebenefratelli Sacco, Luigi Sacco Hospital, Polo Universitario, University of Milan, Milan

85. Infectious Disease Unit, Hospital of Lucca, Italy.

86. Department of Diagnostic and Laboratory Medicine, Institute of Biochemistry and Clinical Biochemistry, Fondazione Policlinico Universitario A. Gemelli IRCCS, Catholic University of the Sacred Heart, Rome, Italy.

87. Clinic of Infectious Diseases, Catholic University of the Sacred Heart, Rome, Italy.

88. Laboratory of Regulatory and Functional Genomics, Fondazione IRCCS Casa Sollievo della Sofferenza, San Giovanni Rotondo (Foggia), Italy

